# Coordinated support for local action: A modeling study of strategies to facilitate behavior adoption in urban poor communities of Liberia for sustained COVID-19 suppression

**DOI:** 10.1101/2020.08.11.20172031

**Authors:** Laura A. Skrip, Mosoka P. Fallah, Jamie Bedson, Laurent Hébert-Dufresne, Benjamin M. Althouse

## Abstract

**Background:** Long-term suppression of SARS-CoV-2 transmission will require context-specific strategies that recognize the heterogeneous capacity of communities to undertake public health recommendations, particularly due to limited access to food, sanitation facilities, and physical space required for self-quarantine or isolation. We highlight the epidemiological impact of barriers to adoption of public health recommendations by urban slum populations in low- and middle-income countries (LMICs) and the potential role of community-based initiatives to coordinate efforts that support cases and high-risk contacts.

**Methods:** Daily case updates published by the National Public Health Institute of Liberia were used to inform a stratified stochastic compartmental model representing transmission of SARS-CoV-2 in two subpopulations (urban poor versus less socioeconomically vulnerable) of Montserrado County, Liberia. Differential transmission was considered at levels of the subpopulation, household versus community, and events *(i.e*., funerals). Adoption of home-isolation behavior was assumed to be related to the proportion of each subpopulation residing in housing units with multiple rooms, access to sanitation facilities, and access to basic goods like water and food. Percentage reductions in cumulative infection counts, cumulative counts of severe cases, and maximum daily infection counts for each subpopulation were evaluated across intervention scenarios that included symptom-triggered, community-driven efforts to support high-risk contacts and confirmed cases in self-isolation following the scheduled lifting of the state of emergency.

**Results:** Modeled outbreaks for the *status quo* scenario differed between the two subpopulations, with increased overall infection burden but decreased numbers of severe cases in the urban poor subpopulation relative to the less socioeconomically vulnerable population after 180 days post-introduction into Liberia. With more proactive self-isolation by mildly symptomatic individuals after lifting of the public health emergency, median reductions in cumulative infections, severe cases, and maximum daily incidence were 7.6% (IQR: 2.2%-20.9%), 7.0% (2.0%-18.5%), and 9.9% (2.5%-31.4%) for cumulative infections, severe cases, and maximum daily incidence, respectively, across epidemiological curve simulations in the urban poor subpopulation and 16.8% (5.5%-29.3%), 15.0% (5.0%-26.4%), and 28.1% (IQR: 9.3%-47.8%) in the less socioeconomically vulnerable population. An increase in the maximum attainable percentage of behavior adoption by the urban slum subpopulation, with the provision of support to facilitate self-isolation or quarantine, was associated with median reductions in cumulative infections, severe cases, and maximum daily incidence were 19.2% (IQR: 10.1%-34.0%), 21.1% (IQR: 13.3%-34.2%), and 26.0% (IQR: 11.5%-48.9%), respectively, relative to the *status quo* scenario.

**Conclusions:** Broadly supported post-lockdown recommendations that prioritize proactively monitoring symptoms, seeking testing and isolating at home by confirmed cases are limited by resource constraints in urban poor communities. Investing in community-based initiatives that determine needs and coordinate needs-based support for self-identified cases and their contacts could provide a more effective, longer-term strategy for suppressing transmission of COVID-19 in settings with prevalent distrust and socioeconomic vulnerabilities.

## Background

During the ongoing novel coronavirus (COVID-19) pandemic, tradeoffs among the feasibility, ethics, and effectiveness of mandatory social distancing have been debated globally and in low and middle-income countries (LMICs) particularly.[1-3] Social distancing measures create socioeconomic constraints for societies and individuals.[4] In turn, the ability to effectively undertake such measures is dependent on access to essential resources, such as food, sanitation facilities, and physical space.

Several countries have developed context-specific adaptations of standard approaches to control,[5] from harnessing the benefits of coordinated public-private partnerships[6] to promoting food delivery as a means of reducing congestion in marketplaces to conducting online funerals.[7] In parallel, independent efforts are being organized to support vulnerable populations during periods of lockdown, with public and private organizations undertaking distribution of basic goods at different scales *(e.g*., [8-11]) A coordinated approach across response pillars to identify, inform, and support presumptive cases in adopting transmission-reducing behaviors could provide one strategy contributing to long-term suppression of the epidemic curves in LMIC contexts without requiring population-level lockdowns.

As part of COVID-19-specific surveillance, countries are innovating approaches for passive and active case detection.[12-14] For the latter, rapid assessment systems (RAS) have been introduced as a means for self-determination of risk and then communication of guidance so that individuals can seek care and undertake personal protective measures to prevent transmission.[13,15] With increasing specificity and sensitivity of symptom-based case definitions for COVID-19,[16,17] such tools hold potential for effectively and efficiently signaling deployment of testing and contact tracing teams. Importantly, notification via RAS would also allow for allocation of goods to support adoption of home quarantine (among high-risk contacts) or isolation (for confirmed, mildly symptomatic or presymptomatic cases). For instance, provision of curtains, masks, food, water, and chamber toilets could support individuals in socioeconomically vulnerable populations, particularly urban slum communities, to participate in recommendations for home-based social distancing and care.

As a long-term strategy that would persist after lifting large-scale lockdowns or states of emergency, such coordination and resources could be managed in partnership with communities. Community-driven surveillance efforts were effective in identifying Ebola transmission clusters during the outbreak in West Africa.[18,19] Additionally, local action had far-reaching and sustained implications for behavior change.[20,21] With rising stigma[22] against COVID-19 survivors and even against those who are taking precautions, such as wearing face masks, local ownership of response efforts will be critical for effective scale-up. Community-based initiatives, with support from the national response, are more likely to reduce disparities in detection and resource allocation across socioeconomically diverse subpopulations by addressing barriers such as resistance due to differential distrust of authorities.[23] They could also lead to innovative, context-specific solutions to supplement symptom-based surveillance, particularly in urban slum settings with high transmission potential yet significantly younger populations than less socioeconomically vulnerable settings.

Notably, a potential tradeoff of symptom-triggered resource deployment relates to the interplay across demographic structures in slum communities, the demographic correlates of severe COVID-19 disease that would signal response, and the reduced health access in slum communities relative to areas of higher SES. In particular, selective mortality among slum-dwelling adults, combined with trends in out-migration among older adults, has been associated with a higher proportion of females in the 50+ slum-dwelling population in sub-Saharan Africa (SSA).[24] Given the age-dependent gender ratio for severe COVID-19 disease[25,26] -- the disease state which most often triggers care-seeking[27] and, in turn, response activities -- the concentration of the population in younger age groups and higher proportion female among old age groups could lead to significant undetected, and therefore unsupported, asymptomatic or mildly symptomatic disease in slum communities. This could be further compounded by the higher overall disease burden in slum communities, as mild disease may not be recognized as COVID-19 or perceived as an issue warranting action.[28]

Here we present a dynamic transmission modeling framework, parameterized for the SSA context, to evaluate longer-term strategies for case detection and intervention after the lifting of lockdowns. We account for heterogeneous impact of contact tracing and home isolation strategies across settings with varying household-level access to food, water, sanitation, and required space as well as varying detection due to differential proportions symptomatic cases with severe disease across urban subpopulations. To the best of our knowledge, this is the first model to incorporate community-level behavior change with disease transmission and to quantify how disparate behavior adoption due to urban poverty leads to disparate gains from response measures during outbreaks. While parameterized with COVID-19-specific epidemiological and response data, the model incorporates data collected through response efforts during the Ebola outbreak in Montserrado County, Liberia, and the proposed strategies reflect lessons learned from efforts during the Ebola outbreak aimed at facilitating mitigation efforts across socioeconomically diverse settings.

## Methods

### Model overview

A stochastic continuous time compartmental model was developed to investigate the impact of interventions addressing barriers to adoption of home-based quarantine and isolation in informal urban communities (Supplementary Material Figure). The base SEIR transmission model was stratified into two subpopulations to represent separate dynamics for urban poor *(i.e*., slum communities) and less socioeconomically vulnerable communities. We considered informal settlements in Liberia *(i.e*., West Point, New Kru Town, Clara Town, Logan Town, Jallah, Slipway, and Peace Island, Supplementary Figure 2) as constituting the urban slum subpopulation based on the definition outlined by Monrovia City Corporation’s Slum Initiative [29] and previous work on urban poor communities in Monrovia.[30] These communities are characterized by overcrowding, availability but limited accessibility to electricity and water, limited sanitation, and poor housing; using data from the most recent census and correcting for underrepresentation[29,31], our urban poor subpopulation constituted approximately 20% of the county’s population (Supplementary Material Methods). Drawing from data and observations during the Ebola outbreak in Liberia, cases living in slum communities were expected to have greater numbers of contacts, higher transmission rates due to nature of contacts amidst overcrowding, and to seek care after longer delays than those in less socioeconomically vulnerable settings (Distributions in Supplementary Material Tables 1-2).[30] In addition, based on gender-specific trends in aging, out-migration, and mortality in urban slum communities,[24] differential detection and proportions of symptomatic cases with severe disease were assumed between the subpopulations, as described below. Slum communities in Liberia are also characterized by a strong network of informal leaders who held roles in supervising active case finding efforts, conducted by peer-nominated community members and in coordination with national surveillance efforts, during the Ebola outbreak.[32] This network was reactivated during the COVID-19 outbreak.

Initial cases were modeled as importations into the less socioeconomically vulnerable subpopulation. For each time step from the first known introduction on March 16 until the airport was closed on April 10, infection importation occurred with a randomly drawn probability to account for possible infections from air travel passengers after discharge from institutional quarantine. Introduction into the seven urban slum communities was assumed to occur after some delay and via either contact between the two subpopulations or, after initial seeding, between urban slum communities within the subpopulation (More details in the Supplementary Material Methods), until all communities had cases and local transmission at the subpopulation-level dominated.

Within each subpopulation, susceptible individuals in the community were infected according to the force of infection, *λ(t*), proportional to the prevalence of infectious individuals in the population and accounting for contact probabilities within and between subpopulations, as well as the relative infectiousness of asymptomatic *(E_iM_*, 1/1.6 times as infectious[33]) and severely symptomatic *(I_is_*, 1.1 times as infectious[34]) individuals compared to mildly symptomatic individuals *(I_iM_*) (Eq. 1, Supplementary Material Table 1). Household members of new infections, move to a separate susceptible compartment with a higher rate of infection derived from a study of household contacts of cases in China (*λ_H_*, Supplementary Material Table 1).[35,36] We assumed a Poisson distribution of individuals per household, such that households of new infections were represented as the average number of individuals per household in Liberia (4.9 individuals[37]) times the number of new infections in the community.

A percentage of all infections remain asymptomatic and the rest develop symptoms at a rate reflecting the incubation period. For the latter, we considered two scenarios. Based on evidence available from high transmission settings, we assumed that most cases with any symptoms in the less socioeconomically vulnerable subpopulation remained mildly symptomatic (1 − *s*),[38,39] while 19% were assumed to develop severe symptoms *(s* = 0.19). In addition, to account for differing age structures and underlying comorbidities in West Africa, we ran the model to reflect the scenario with a lower percentage, 15%, of symptomatic individuals assumed to develop severe symptoms.[40] For the urban slum subpopulation, we assumed that the percentage of symptomatic cases with severe symptoms would be reduced by a factor equal to the ratio of crude death rates among 50+ y.o. individuals in urban slums versus the entire population, as determined by a study in Nairobi *(i.e*., 22.8/40.9 × *s)*. Those cases with severe symptoms had a rate of disease-associated death *(π)*. Mild cases and severe cases who survive the virus recover at a rate *ω*. Recovered individuals remained immune, in the absence of clear evidence about duration of protection.[41]

For the base model reflecting the *status quo* situation of response efforts to date in Liberia, quarantine and isolation were undertaken by some of the individuals identified through contact tracing. The impact of contact tracing on transmission was modeled by having a subset of new infections immediately removed from the infectious population; that is, a percentage *(g)* of pre-symptomatic individuals would undertake quarantine and remain isolated through recovery or death, either at home or in a hospital depending on severity of symptoms, due to contact tracing. The impact of contact tracing was assumed to be time-varying and account for both the effectiveness of intervention in accessing contacts that ultimately became cases as well as the level of distrust or denial in the population around COVID-19 which was assumed to affect willingness to isolate among those traced (More details in Supplementary Material). Additionally, among cases who develop severe symptoms but were not listed as contacts, a time-varying percentage (*e*_2_), assumed to be maximum 30% based on observations in care-seeking during the Ebola outbreak, undergo isolation in a hospital at a rate (*τ_i_*) or may remain in the community. Hospitalization removed individuals from the infectious population and home-isolation reduced the infectiousness of individuals by 80% to reflect estimates for the effectiveness of mask-wearing.[42] Rates of isolation (*τ_i_*) were drawn from subpopulation-specific distributions reflecting days to care-seeking during the Ebola outbreak [30] (and consistent with observations for care-seeking after onset of COVID-19 in China[43]) and percentages of isolation adoption (*e*_1_) were subpopulation-specific based on access in resources to do so.

It is expected that realities around overcrowding and shared facilities may inhibit the ability to adopt home isolation recommendations with fidelity, regardless of willingness to do so. Therefore, the percentage of cases adopting home-isolation behavior was assumed to be related to the proportion of the subpopulation residing in housing units with multiple rooms, access to sanitation facilities, and access to basic goods like water and food (Supplementary Material Table 2). These factors would differentially affect the two subpopulations in the model. Data on needs around water, food, and toilet access in informal urban settlements versus less socioeconomically vulnerable urban communities were derived from surveys conducted in Kenya.[44,45] In the less socioeconomically vulnerable urban communities, adoption of home quarantine or isolation would be undertaken by the percentage of respondents from Liberia who had indicated that it was very easy, easy, or neither easy nor difficult to adhere to social distancing on a crowdsourcing survey,[46] as respondents were assumed to be more likely representative of this subpopulation than the urban poor subpopulation.

A separate compartment for community deaths *(i.e*., deceased cases who did not seek care) was used to incorporate transmission clusters due to funerals, as has been observed for SARS-CoV-2 due to the aggregation of high numbers of attendees.[47,48] A number of contacts was drawn for each community death based on the number of individuals in attendance at unsafe burials *(i.e*., traditional burials occurring after recommendations around safe burials were in place) reported in West Africa during the Ebola outbreak and were infected at a rate equal to *λ_F_ = β_i_ × I_it_/N_it_* for *I_it_/N_it_* representing the prevalence of infection at each time *t* for subpopulation *i*. The distribution of traditional burial attendees for community deaths reflects data from after policies were put into effect for safe burials and therefore were expected to reflect the present case in Liberia despite policies limiting funeral attendance to 10 individuals. The Liberian government also instituted policies to reduce numbers of attendees at religious gatherings, with public support from religious leaders and the Council of Churches [49], and at sporting events. We assumed that institution-level, organized events would have greater enforcement of lockdown-related policies and did not explicitly include those in the model.

### Model fitting

An expression for *R_o_* was derived using the next generation matrix approach[50] assuming introduction and initial spread in the less socioeconomically vulnerable subpopulation was calibrated to achieve an average subpopulation-level *R_o_* of 2.4 (SD: 0.2). A likelihood-based approach was used to compare model output across sets of parameters, including the transmission parameter *β*, calibrated using NGM to cumulative case data from Montserrado County, Liberia (Supplementary Material Methods). Lastly, Bayesian melding[51] was implemented to probabilistically sample 1,000 parameter sets according to the weight of their normalized negative log likelihood values for subsequent model projections to evaluate intervention scenarios.

### Intervention scenarios

Intervention results were evaluated across both assumptions for *s* = 0.15,0.19. For all intervention scenarios, we ran the *status quo* model until the end of the state of emergency and partial lockdown slated for July 21 (after a 30-day extension was declared by the President on June 22[52]), or 128 days post-introduction into Liberia, and then introduced the intervention with scale-up for one month (according to a step function with a uniform percentage change every 5 days), followed by maximum coverage for the subsequent three weeks. The *status quo* scenario was based around care-seeking by severely symptomatic individuals or isolation of high-risk contacts due to top-down initiatives through contact tracing. We evaluated the difference in cumulative incidence and maximum daily incidence over a 180-day (approximately, six months from introduction) period between each curve from the *status quo* scenario and the curve with corresponding parameter set for each intervention scenario.

Under the intervention scenario of *Self-identification without support*, we investigated how community-level efforts with self-identification, such as through RAS, by individuals not on contact tracing lists and with mild symptoms would lead to greater and more proactive coverage of self-isolation by community members. Home isolation by mildly symptomatic individuals would be limited by the subpopulation-specific maximum attainable percentage based on resource constraints.

Under the intervention scenario of *Self-identification with community-driven support*, we modeled coordinated efforts to provide food, water, chamber toilets, masks and other materials necessary for undertaking home-based quarantine or isolation. This was incorporated as an increase in the percentage of adoption (e_1_) in the urban poor subpopulation. We assumed that the provision of these materials was coordinated through community engagement efforts that were also associated with improved overall adoption of public health recommendations.[20] This was modeled across varying reductions in the rates of isolation and treatment-seeking and in the number of contacts attending funerals. Specifically, we explored the role of support in reducing rates of isolation and number of funeral attendees by 25%, 50%, and 75%.

Thus, without support, the maximum attainable percentage of the urban poor subpopulation adopting home isolation or quarantine was determined by the percentage of the subpopulation having resources to do so, namely, access to sanitation, food, water, and physical space needed to safely and effectively do so. With support, the percentage adoption was modeled to reflect equal adoption across the entire population.

## Results

Given uncertainty in actual infection rates due to likely underreporting, a range of probable epidemic curves, accounting for social distancing *(i.e*., partial lockdown and school closures) and contact tracing measures in place, were considered across intervention scenarios (Figure 1). Accounting for differential transmission probabilities, contact numbers, mechanisms and timing of infection introduction, and proportion of severe cases, differently shaped and timed epidemic curves were observed for the two subpopulations (Figure 2).

**Figure 1.**
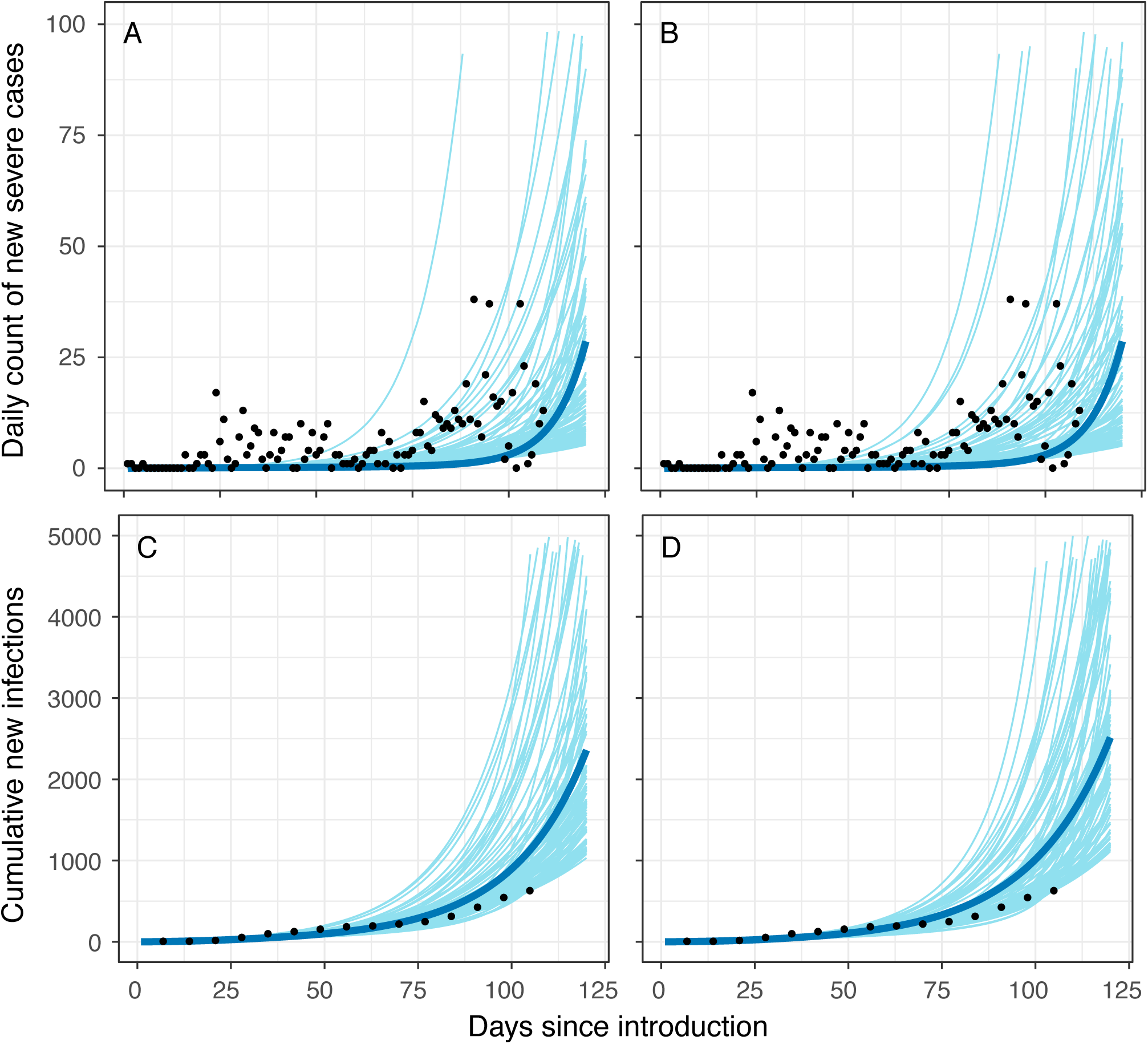
Modeled daily incidence of severe cases and cumulative infection counts in Montserrado County for assumptions of (A,C) 19% severity among symptomatic cases and (B,D) 15% severity among symptomatic cases. The blue line represents the median of all curves selected via the Bayesian melding approach, conducted separately for each assumption of severity percentage. Gray curves represent 100 random draws from parameter sets selected via Bayesian melding. Black dots represent data points (daily reported cases or cumulative reported cases at weekly intervals) per reports of confirmed cases from the National Public Health Institute of Liberia.

**Figure 2.**
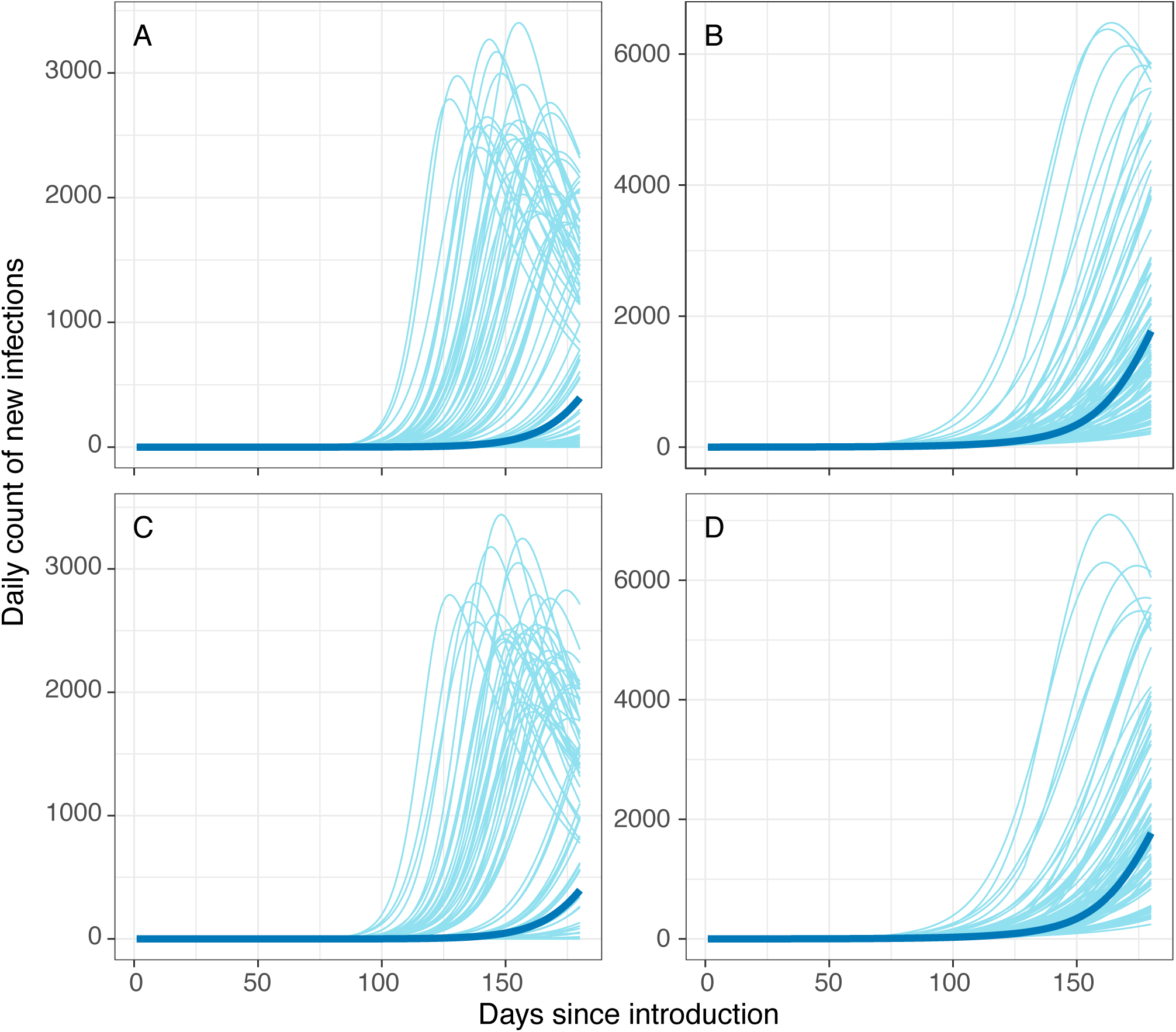
Pre-intervention epidemic curves in the (A, C) urban poor subpopulation and (B, D) less socioeconomically vulnerable subpopulation. Gray lines represent 100 random draws from the curves that have 150-day cumulative infection counts in the interquartile range. Blue lines represent curves that are associated with median 150-day cumulative infection counts. Top row panels (A-B) reflect results from assumption of 19% severity among symptomatic cases and bottom row panels (C-D) reflect results from assumption of 15% severity among symptomatic cases.

Assuming 19% severity among symptomatic cases in the less socioeconomically vulnerable population, simulations of a 180-day (approximately, six-month) outbreak were associated with a median cumulative 6,005 infections (IQR: 105-88,217) and a median maximum daily incidence of 21 per 10,000 people (IQR: <1-121) in the subpopulation of urban poor; a median cumulative 35,926 infections (IQR: 19,792-65,569) and median maximum daily incidence of 17 per 10,000 people (IQR: 9-29) were estimated for the less socioeconomically vulnerable subpopulation -- a 25% difference in incidence (Figure 2). With an assumption of 15% severity among symptomatic cases, a median cumulative 6996 infections (IQR: 114-90,324) and a median maximum daily incidence of 27 per 10,000 people (IQR: <1-121) were observed after 150 days in the subpopulation of urban poor; a median cumulative 37,304 infections (IQR: 19,259-64,318) and median maximum daily incidence of 18 per 10,000 people (IQR: 9-30) were estimated for the less socioeconomically vulnerable subpopulation -- a 50% difference in incidence (Figure 2).

We evaluated reductions in incidence due to the introduction of home isolation of mild cases, such as due to self-protective measures in response to feedback from use of RAS or local action by families and communities in reaction to symptoms (Figures 3-4). This was introduced after the lockdown ended (July 21) and was implemented under the assumptions that severely sick individuals continue to seek treatment in a hospital at rates consistent with those prior to the end of the lockdown and that contact tracing efficiency and effectiveness would likewise remain consistent. In general, burden averted in the less socioeconomically vulnerable subpopulation was as much as three times greater than that in the urban poor subpopulation.

**Figure 3.**
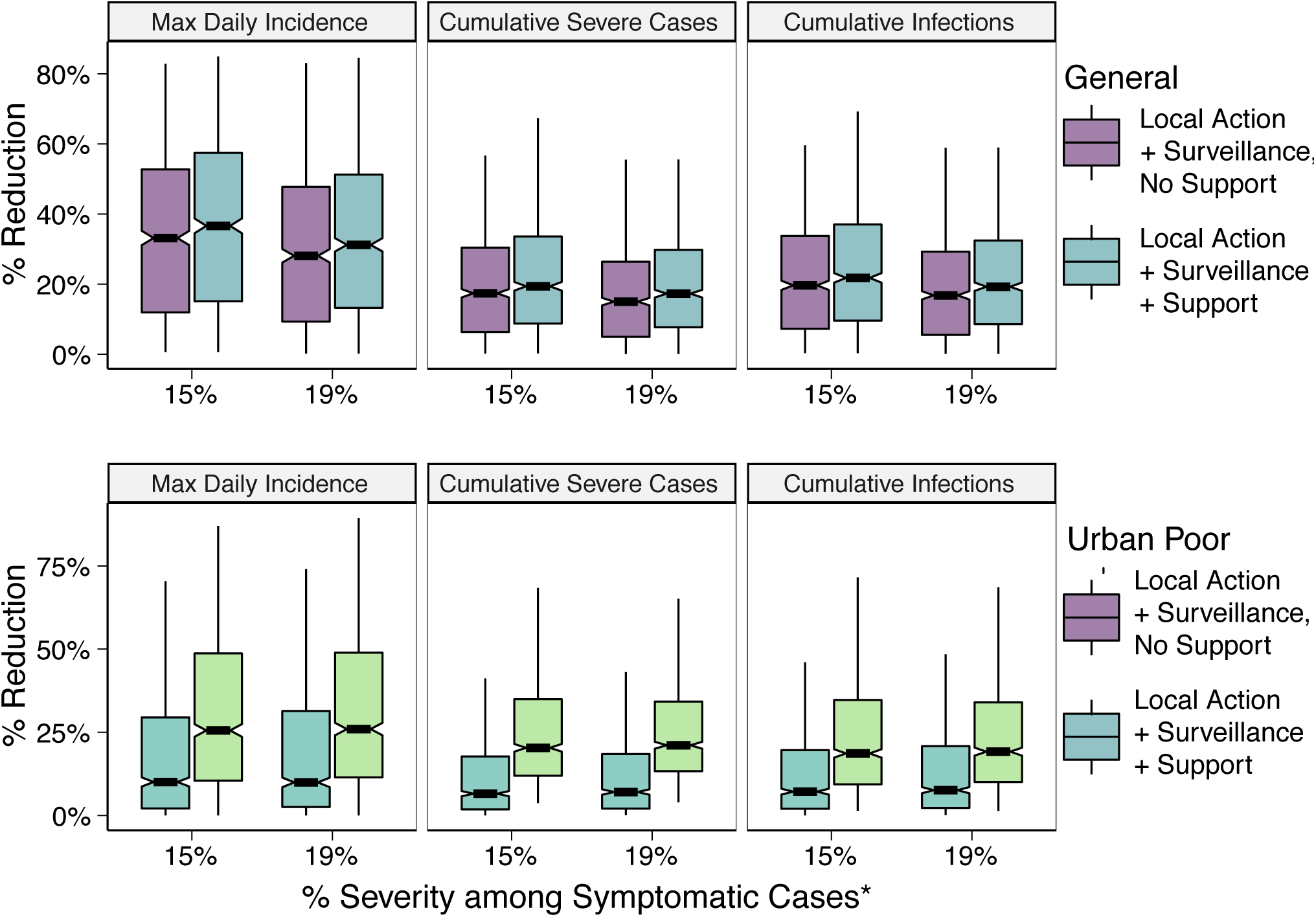
Percentage change in cumulative infections, cumulative severe cases, and maximum daily incidence (peak of the curve) with post-lockdown introduction of a symptom-based self-identification intervention, relative to continuation of status quo interventions alone. Subpopulation-specific impact of interventions with and without community-coordinated support efforts are presented. *Results are presented for different assumptions around relative proportions of severe versus mild symptomatic cases in the less socioeconomically vulnerable subpopulation (labeled “General population”).

**Figure 4.**
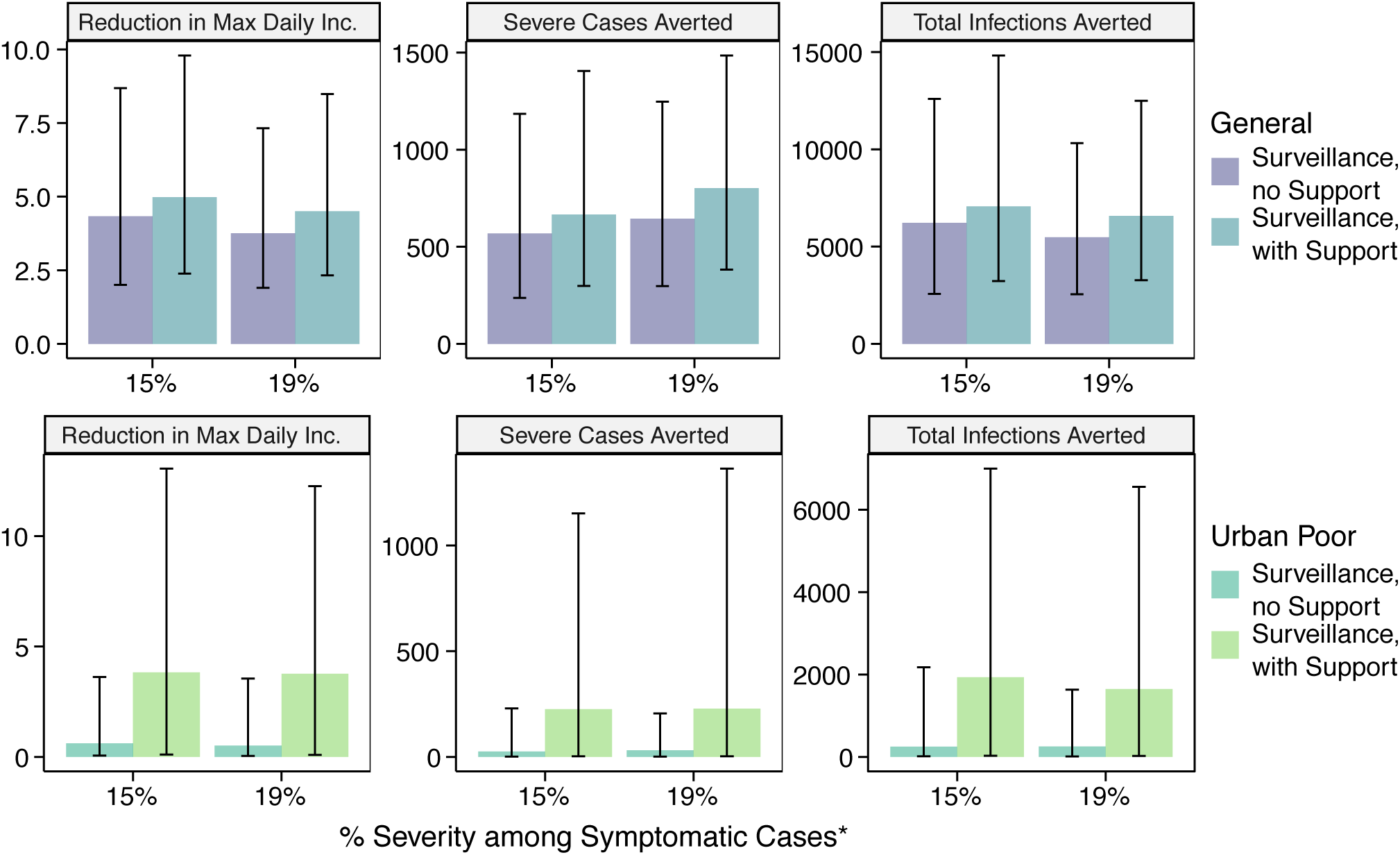
Median change in cumulative infections, cumulative severe cases, and maximum daily incidence (peak of the curve) with post-lockdown introduction of a symptom-based self-identification intervention, relative to continuation of status quo interventions alone. Subpopulation-specific impact of interventions with and without community-coordinated support efforts are presented. *Results are presented for different assumptions around relative proportions of severe versus mild symptomatic cases in the less socioeconomically vulnerable subpopulation (labeled “General population”). Error bars represent interquartile range.

For the assumption of 19% symptom severity, adoption of home isolation up to the maximum attainable percentage without support was associated with median reductions of 7.6% (IQR: 2.2%-20.9%), 7.0% (2.0%-18.5%), and 9.9% (IQR: 2.5%-31.4%) cumulative infections, severe cases, and maximum daily incidence, respectively, in the urban poor subpopulation (Figure 3). This reflects a median 256 infections (IQR: 12-1,635) and 32 (IQR: 1-206) severe cases averted, relative to the *status quo* scenario (Figure 4). In the less socioeconomically vulnerable subpopulation, median reductions were 16.8% (IQR: 5.5%-29.3%), 15.0% (5.0%-26.4%), and 28.1% (IQR: 9.3%-47.8%) for cumulative infections, severe cases, and maximum daily incidence. This reflects a median 5,479 (IQR: 2,548-10,321) infections and 645 (IQR: 298-1,248) severe cases averted, relative to the *status quo* scenario. For the assumption of 15% symptom severity, introduction of home isolation of mild cases without support was associated with 7.2% (2.0%-19.6%), 6.5% (IQR: 1.8%-17.7%), and 10.1% (IQR: 2.1%-29.5%) reductions in total infections, severe cases, and maximum daily incidence in the urban poor subpopulation and 19.7% (IQR: 7.3%-33.8%), 17.4% (IQR: 6.4%-30.4%), and 33.2% (IQR: 12.0%-52.7%) reductions in the less socioeconomically vulnerable subpopulation, relative to the *status quo* scenario. All results for both assumptions around symptom severity are included in Supplementary Tables 3-4.

For the urban poor subpopulation, the most significant increase in impact was observed with the introduction of support, by which the proportion of the subpopulation with necessary resources to safely and effectively quarantine or self-isolate was increased to that of the less socioeconomically vulnerable subpopulation. Median reductions in cumulative infections, severe cases, and maximum daily incidence were 19.2% (IQR: 10.1%-34.0%), 17.3% (IQR: 7.7%-29.8%), and 26.0% (IQR: 11.5%-48.9%), respectively, relative to the *status quo* scenario, under the assumption of 19% symptom severity for the less socioeconomically vulnerable and 18.6% (IQR: 9.4%-34.7%), 20.3% (IQR: 11.9%-35.0%), and 25.5% (10.5%-48.7%), respectively, under the assumption of 15% symptom severity (Figure 3). This corresponds to a median 1,650 infections (IQR: 24-6,557) and 228 (IQR: 3-1,363) severe cases averted relative to the *status quo* scenario for 19% symptom severity and 1,933 (IQR: 30-6,998) infections averted and 226 (IQR: 3-1,152) severe cases averted for 15% symptom severity. Additional impact was observed with simultaneous reductions in time to isolation and/or number of funeral contacts across all symptomatic cases (Figure 5). However, even with support, 75% reduction in time to isolation, and 75% reduction in funeral contacts, transmission in the urban poor population was sustained at low levels. A median 61.1%% (IQR: 47.1%-77.2%) of infections from the unmitigated scenario would be expected in this scenario at 180-day post introduction, assuming 19% symptom severity.

**Figure 5.**
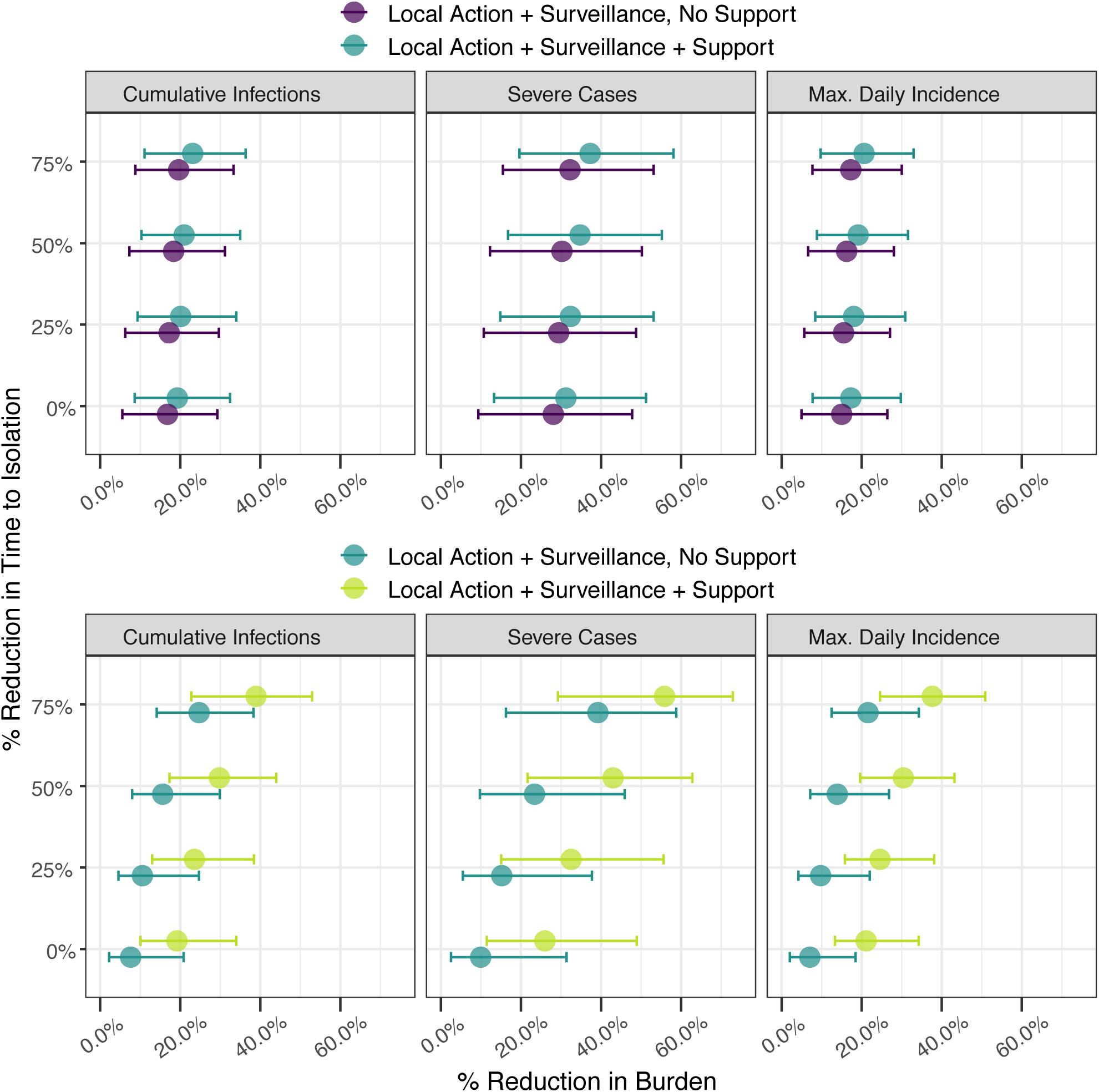
Percentage reduction in infections, cases of severe disease, and maximum daily incidence in the less socioeconomically vulnerable subpopulation (top panel, blue color) and urban poor subpopulation (bottom panel, green color) with introduction of efforts to suppress COVID-19 transmission. Interventions including contact tracing and surveillance efforts were evaluated across simultaneous reductions in time to isolation. Results reflect the assumption of 19% symptom severity.

For the less socioeconomically vulnerable subpopulation, the provision of support for the urban poor subpopulation was associated with further transmission reduction, although the added benefit of surveillance alone, relative to local action only, was more drastic than that of surveillance and support, relative to surveillance alone. Median percent reductions in cumulative infections, severe cases, and maximum daily incidence were 19.3% (IQR: 8.6%-32.5%), 17.3% (IQR: 7.7%-29.8%), and 31.2% (IQR: 13.3%-51.2%), respectively, relative to the *status quo* scenario, under the assumption of 19% symptom severity. Reductions of 21.8% (IQR: 9.6%-37.0%) in cumulative infections, 19.4% (iQr: 8.8%-33.6%) in severe cases, and 36.6% (IQR: 15.1%-57.4%) were observed under the assumption of 15% symptom severity. These reductions correspond to averting 7,069 (IQR: 3,230-14,818) infections and 665 (IQR: 299-1,405) severe cases relative to the *status quo* for the 19% severity assumption and averting 6,575 (IQR: 3,275-12,494) infections and 802 (IQR: 382-1,485) severe cases for the assumption of 15% symptom severity.

## Discussion

We developed a stratified compartmental modeling framework to consider adoption of home isolation and quarantine across socioeconomically diverse urban subpopulations. Despite later introduction into the urban poor subpopulation and less severity of disease due to sociodemographic characteristics, our assumptions around relative probability of transmission between subpopulations as well as differential contact and other behavior patterns suggest the potential for a more severe peak in the urban poor communities unless contact tracing at the current levels of effectiveness and efficiency are supplemented with alternative strategies post-lockdown. Our results highlight how reducing heterogeneity in behavior adoption by supporting high-risk contacts and mildly symptomatic or asymptomatic individuals in urban poor areas to undertake recommendations could lead to nearly a 20% reduction in both infections and severe cases, relative to the *status quo* approach, in that subpopulation, compared to less than 10% reductions in the absence of support. Additional gains due to quicker care-seeking would be expected with well-coordinated support efforts that built trust through community-led response. To mitigate burden in the urban slum subpopulation and overall, rapid and effective intervention among high risk contacts is important; it is expected that, per our model, that delays in introducing additional interventions post-lockdown could result in the numbers of contacts requiring more resource-intensive support-based surveillance efforts overwhelming even locally based efforts. Low-cost, universal efforts at prevention *(i.e*., to reduce transmission probability), such as through personal hygiene and use of masks, would therefore be important to reduce the excess burden in urban poor communities as well as enhance the feasibility of surveillance and support strategies.

We have incorporated model adaptations to reflect the Liberia-specific response and the context of West Africa in general, including potentially lower percentage of disease severity among cases, the role of funerals as high-transmission events due to close congregation of individuals, and the impact of constraints around food, water, physical space, and sanitation on adoption of social distancing interventions. Previous work has investigated how the sociodemographic characteristics of urban slum communities influence disease transmission dynamics.[53] These were implemented to consider the impact of community-initiated, coordinated efforts across response pillars for identifying and supporting cases and their high-risk contacts in order to suppress the epidemic curve without population-level lockdown. The adaptations reflected in this framework have relevance for ongoing transmission in settings with overcrowding and resource constraints--such as with the recent introduction of SARS-CoV-2 into UN camps in South Sudan[54] and ongoing reports of outbreaks in prisons[55] homeless shelters, and under-resourced aged care homes[56], globally. They also are pertinent to settings with socioeconomic disparity, with the goal of promoting consideration of approaches that engender local agency and facilitate adoption of transmission-reducing practices in complex settings.

We evaluated both surveillance strategies, with and without support, across different changes in time to isolation and numbers of attendees at funerals for community deaths. It is expected that such behavioral changes would be more likely with a coordinated, community-led effort in which people felt supported. Community-based initiatives that contributed to surveillance have precedent in controlling infectious disease outbreaks [21,32] and will be essential to longer term efforts during the COVID-19 pandemic, particularly as funds for government-led, large-scale programs may not sustain top-driven efforts at contact tracing. In the context of COVID-19, community engagement in slum communities would entail i) well-designed and supported community-led surveillance that is integrated into the overall response; ii) development and monitoring of feedback mechanisms *(i.e*., goods distribution, symptom monitoring, testing, etc.) that encourage and reinforce self-isolation of mildly symptomatic, symptomatic and presymptomatic cases; and iii) development and dissemination of messaging in collaboration with communities that addresses the complexities of these strategies (i.e. why a presymptomatic person needs to self-isolate). Such a strategy could begin to address undetected transmission in marginalized urban communities and across the overall population and could reduce the need for prolonged states of emergency or other top-down approaches with social and economic consequences.

National coordinated resource allocation in HMICs and LMICs has primarily focused on distribution of testing materials, procurement of ventilators, and maintenance of isolation centers. While such efforts are critical for improving detection of cases and reducing mortality among severe cases, they tend to overlook strategies for mildly symptomatic, asymptomatic, and presymptomatic cases that could reduce overall transmission and protect individuals at highest risk of severe disease. This may be due to assumptions that people will (and have the resources needed to) undertake home-based self-care, wash hands, and wear masks, or due to policies to isolate all positive cases in facilities. Our findings indicated that symptomatic surveillance alone has limited impact on reducing overall transmission, particularly in the absence of support to individuals in home isolation. However, the use of self-reported symptoms to trigger responsive, community-engaged efforts that facilitate home isolation and other protective measures could offer a scalable, lower cost policy which can be embedded within a larger, multi-pronged response strategy that does not depend on mass lockdowns. It is important to emphasize that, despite challenges, innovations for reducing transmission in complex settings exist (e.g., [57,58]) and evaluation of their individual or combined potential is important. Such innovations go beyond the standard non-pharmaceutical interventions being modeled [59,60] for HMIC and LMIC settings and may warrant investments, such as in sustainable infrastructure and development projects, with longer term impacts.[61]

Like all studies, ours is not without limitations. The current model draws on information from varied sources around behavior change during past outbreaks and conditions in LMIC urban settings outside of Liberia. We recognize that the clinical presentation of SARS-CoV-2 infection may result in different behavior with and between subpopulations than Ebola virus disease. The model is therefore intended to offer a framework that can be used to explore country-specific policies with locally available data, or proxy information in the absence of data, on heterogeneous socioeconomic and demographic characteristics of urban populations and COVID-19-specific dynamics. As such, data-driven applications of the framework fit to subpopulation-specific and age-stratified data on infection rates, the ratio of mildly to severely symptomatic cases, and adoption of home-isolation behaviors in urban settings will be critical next steps. Such data will also allow for the model to directly or indirectly account for other barriers to adoption, including fear of stigma, constraints around surveillance efforts, low overall rates of testing due to capacity constraints at the national level, and differences in testing between subpopulations. Recognizing the complexity and interconnectedness of factors contributing to challenges around health access and outcomes in slum communities, COVID-19-specific surveys on barriers to adoption of public health recommendations would improve the coarse assumptions made in the present model. Furthermore, we fit the model to data from the first 105 days post-introduction in Liberia, and as has been experienced in countries globally, low testing rates and data unavailability lead to uncertainty around the extent of the epidemic. For instance, the country has been implementing school closures and a partial lockdown, which are affecting contact patterns in currently unmeasured ways and which may be suppressing the curve more than our *status quo* results suggest.

It is important to highlight the potential of low-cost and long-term innovations that will support individuals and communities to undertake transmission-reducing behaviors and actively participate in the greater response. While organizations are independently providing food, water, and other items to address basic needs, a coordinated effort that directs resources based on well-designed and supported community-led epidemiological surveillance findings could more effectively overcome barriers to adoption of public health recommendations, particularly in urban slum communities. Importantly, investing in community-driven initiatives that extend impact from resource allocation to heightened sense of agency, such as that which would encourage more and quicker treatment seeking and adoption of safe burial practices, could lead to significant additional reductions in case counts and deaths. Integration of such efforts into a national mitigation strategy could lead to greater reductions in infection rates and numbers of severe cases than less community-engaged efforts.

## Data Availability

Parameters used for the model are included in Supplementary Tables 1 and 2 and, where appropriate, referenced. Data used for fitting the model are from publicly available sources, namely the daily updates prepared by the National Public Health Institute of Liberia and published on its Facebook page:
https://web.facebook.com/National-Public-Health-Institute-of-Liberia-NPHIL-164280647325112/?_rdc=2&_rdr

